# Demographic variation and socioeconomic inequalities associated with the triple burden of malnutrition in Vietnamese children aged 6 months to 9 years old: Findings from the Vietnamese General Nutrition Survey 2020

**DOI:** 10.1101/2024.03.18.24304456

**Authors:** P.Y. Tan, V. S. Som, S. D. Nguyen, X. Tan, D.T. Tran, T.N. Tran, V.K. Tran, L. Dye, J. B. Moore, S. Caton, H. Ensaff, X. Lin, G. Smith, Y. Y. Gong

## Abstract

**Introduction:** The triple burden of malnutrition (TBM) is a growing public health issue worldwide. This study examined the prevalence and association between undernutrition, overnutrition and micronutrient deficiencies (MNDs), and the associated demographic and socioeconomic determinants, among Vietnamese children, using the nationally representative General Nutrition Survey 2020.

**Methods:** Data on anthropometric parameters, micronutrients biomarkers, demographic and socioeconomic indicators for 7,289 children aged 6 months to 9 years old were analysed. Determinants of malnutrition were assessed using logistic regressions and reported as odds ratio (OR) [95% confidence intervals (CI)].

**Results:** Overall, 12.7%, 10.5% and 4.7% of children were stunted, underweight and wasted/thin; while 7.3% and 7.1% were living with overweight and obesity, respectively. Low serum zinc, anaemia and iron deficiency (ID) were the common MNDs observed, affecting 53.1%, 15.2% and 13.9% of the study participants. Older children aged 2-4 years old [OR (95% CI): 1.43 (1.20, 1.72)], ethnic minorities [5.94 (3.78, 9.36)] and living in mountainous areas [5.06 (1.18, 14.42)] had increased odds of stunting, whereas reduced odds were found in children from the richest quintile [0.13 (0.05, 0.32)]. Similar determinants were found to be associated with underweight and MNDs. Males [1.43 (1.16, 1.76)], children aged 5-9 years old [10.02 (6.71, 14.97) and children from the richest quintile [2.91 (1.20, 7.05)] had increased odds of overweight. Children with anaemia, low serum retinol and low serum zinc had increased odds of stunting and underweight than non-micronutrient deficient children (adjusted OR=1.43-1.71). Compared to children without MNDs, those with ≥3 MNDs had almost double the odds of stunting and underweight, whereas those with ≤3 MNDs had reduced odds of overweight (adjusted OR=0.38-0.60).

**Conclusions:** Significant demographic variation and socioeconomic inequalities in child malnutrition were identified. National policies and programmes in Vietnam should address age-specific, sex-specific, geographical and socioeconomic disparities to accelerate progress in reducing child malnutrition.

**What is already known on this topic?** *summarise the state of scientific knowledge on this subject before you did your study and why this study needed to be done*

- The triple burden of malnutrition (TBM) refers to the co-existence of undernutrition, overnutrition and micronutrient deficiencies (MNDs). Although TBM is a major public health issue that affects every country in the world, it is particularly pervasive in low- and middle-income countries.
- There is no recent data investigating the magnitude of the TBM in Vietnam and how prevalence of TBM differs across different demographic and socioeconomic groups, especially amongst infants and young children.

**What this study adds?** *summarise what we now know as a result of this study that we did not know before*

- Our analysis of the Vietnamese national representative General Nutrition Survey (GNS) 2020 data demonstrated the extent of the co-existence of each aspect of the TBM among children in Vietnam, and demographic variation and socioeconomic inequalities in child malnutrition were identified.
- Although child stunting reduction is on track towards the target of 40% reduction by 2025 set by the Global Nutrition Targets, prevalence is still high at10%-15% in 2020. Meanwhile overweight and obesity in school aged children is off track the target with the prevalence in urban areas increasing from 8.5% in 2010 to 31% in 2020.
- Stunting, underweight and MNDs were more prevalent in infants and young children, particularly in ethnic minorities, living in the rural and mountainous areas and poorer families. Childhood overweight and obesity were evident, especially among school-aged children, males, living in the urban areas and richer families.
- Low serum zinc (53%), anaemia (15%) and iron deficiency (14%) were the most common MNDs, and prevalence of low serum retinol was relatively low (<7%) in this population.

**How this study might affect research, practice or policy?** *summarise the implications of this study*

- These findings emphasise the need for double-duty actions to simultaneously address all forms of child malnutrition in Vietnam.
- National food and nutrition policies and programmes in Vietnam should address age-specific, sex-specific, geographical and socioeconomic disparities to ensure equitable access and opportunities for all, to accelerate progress in reducing child malnutrition.

## Introduction

Malnutrition in all forms, including undernutrition, overnutrition and micronutrient deficiencies (MNDs) currently affects every country in the world to a greater or lesser degree. Child malnutrition exerts significant impacts on a country’s economy as it reduces human productivity and creates a burden on the health care system (1). Undernutrition is known to increase the risk of child mortality and morbidity in children under five, and the associated MNDs such as iron, zinc, folate, iodine and vitamin A increase susceptibility to infections, impair immune function and reduce growth and development (2). Children living with overweight and obesity are more likely to become adults with obesity and an elevated risk of obesity-related non-communicable diseases (NCDs) at a younger age (3, 4). Moreover, life course evidence shows that exposure to early undernutrition is linked to overweight and obesity that later in life will increase the risk of NCDs (5). Thus, childhood is a critical period to address nutrition-related disease burden and prevent intergenerational malnutrition and its consequences.

Vietnam has committed to achieving the targets set in the UN Sustainable Development Goals (SDGs) (6). In particular, SDG 2.2 calls for an end to all forms of malnutrition. By 2030, the Vietnamese government aims to reduce child stunting and underweight in children aged <5 years old to <20% and <10%, respectively, and to reduce the prevalence of overweight in urban areas to <10% (6). The General Nutrition Survey (GNS) reported that the prevalence of stunting in Vietnamese children aged under five has been substantially reduced from 43.3% in 2000 to 29.3% in 2010 and to 24.5% in 2015. The prevalence of underweight has also reduced from 30.1% in 2000 to 17.5% in 2010 and 13.9% in 2015 (6–8). However, the reduction in undernutrition has been slower in the rural areas of the Northern, the South-Central and the Central Highlands regions. In contrast, in urban regions, there has been an increasing trend in childhood obesity. Whereby the prevalence of overweight and obesity among children and adolescents aged 5–19 years has risen from 8.5% in 2010 to 19.0% in 2020, and is higher in urban and rural areas than in mountainous areas (26.8% and 18.3% vs. 6.9%) (9). This regional disparity in undernutrition and overnutrition which is related to the inequalities in socioeconomic growth and development between regions, poses significant challenges for Vietnam to achieve their national goals in ending all forms of malnutrition in the near term (10).

MNDs negatively affect human health over the life course, but have been under-recognised (11). This “hidden hunger” is increasingly evident in individuals with both undernutrition and overnutrition (12). Using pooled analyses from nationally representative population-based surveys spanning 2003-2019, Stevens and colleagues (2022) reported that nearly 56% (∼372 million) of children aged 6-59 months globally were suffering from at least one of the three common MNDs (iron, zinc and vitamin A) (13). In Vietnam, MNDs have been reported to affect nearly 66% of children aged <5 years old, with 56%, 19%, 6% and 14% experiencing iron, zinc, vitamin A and vitamin D deficiencies, respectively (13). Both food insecurity (e.g., energy and nutrient inadequate diets) and micronutrient-poor, energy-dense diets are strongly associated with MNDs (14, 15). This co-occurrence has prompted the term “triple burden of malnutrition” (16).

To date, there are limited recent data on the prevalence and associations between multiple burdens of malnutrition, and relevant demographic and socioeconomic factors in Vietnam, especially among children. Therefore, this study aimed to determine the prevalence of undernutrition (stunting, underweight and wasting/thinness), overnutrition (overweight and obesity) and MNDs (anaemia, iron, vitamin A or retinol and zinc deficiencies), in Vietnamese children aged 6 months to 9 years old, to explore its associations with demographic and socioeconomic indicators, and to examine the relationship between MNDs and undernutrition, and overnutrition, using data from the nationally representative Vietnamese GNS 2020.

## Methods

### Study population and study design

Secondary analysis of the nationally representative Vietnamese GNS 2020 was performed. The GNS is conducted every 10 years by the National Institute of Nutrition (NIN). A multiLJstage cluster sampling technique was used to select a representative sample of Vietnamese population. Clusters corresponded to census enumeration areas (EAs) and were treated as primary sampling units. The first stage of sampling involved selection of a sub-set of provinces across six geographical areas in Vietnam, namely i) Northern midlands and mountainous areas, ii) Central highlands, iii) Red River Delta (including Hanoi Capital); iv) North-central and central coastal areas, v) Southeast and vi) Mekong River Delta (including Ho Chi Minh City). EAs were selected from 25 out of 63 provinces to represent the six geographical areas. In the second stage of sampling, the EAs from the selected provinces were allocated to an urban, rural and mountainous stratum to ensure representation of urbanicity. In the final stage of sampling, eligible individuals from each EAs were randomly selected for enrolment in GNS 2020. Participants encompassed various sub-groups, including those aged 0-5, 6-9, 10-14, and 15-49 years old, along with pregnant and lactating women. For children aged ≤9 years old, informed written consent was obtained from their parents/caregivers. Individuals who declined to respond or did not provide written informed consent were included from the survey. A total of 20,864 individuals were enrolled in GNS 2020. In this study, individuals were excluded if they had missing age data or if they aged <6 months and >9 years old. Finally, a total of 7,829 children aged 6 months to 9 years old who had complete demographic indicators were included for analysis.

### Anthropometric measurements

All anthropometric measurements including body weight and height/length were taken in children wearing light clothes and without shoes. All measurements were taken twice, and the mean was calculated. The anthropometric status of the children was classified according to the WHO Child Growth Standards for children aged <5 years old (17), and WHO Growth Reference for children aged 5-9 years old (18), depending upon the child’s age. Stunting was defined as height-for-age z-scores (HAZ) < −2 standard deviation (SD). Underweight was defined as weight-for-age z-scores (WAZ) < −2 SD. For children aged <5 years old, wasting was defined as weight-for-height z-scores (WHZ) < −2 SD, whereas for children aged 5-9 years old, thinness was defined as body mass index (BMI)-for-age z-scores (BAZ) < −2 SD. BMI was defined as weight (kg) divided by the square of height (m). Overweight and obesity were defined as weight-for-height z-scores (WHZ) > +2 SD to ≤ +3 SD and > +3 SD for children aged <5 years old (17), and BMI-for-age z-scores (BAZ) > +1 SD to ≤ +2 SD and > +2 SD for children aged 5-9 years old (18), respectively. Children with HAZ and WHZ < −6 SD and > +6 SD or children with WAZ < −6 SD or > +5 SD compared to the WHO median value were flagged as outliers and were removed as per WHO recommendation (17, 18).

### Biochemical analysis

Blood samples of children aged 6 months to 9 years old were drawn by venepuncture by experienced nurses following rigorous blood collection, storage and analysis SOPs determined by National Institute of Nutrition (NIN) Vietnam. A total of 5 mL non-fasting blood was collected from each participant. Haemoglobin (Hb) was measured for each participant immediately after blood sample collection using a HemoCue® Hb 301 analyser (HemoCue Angholm, Sweden). The Hb value was recorded and provided to the participant’s parent or caregiver. Any participant who had severe anaemia (<7.0 g/L for children) was referred to the physician at the Commune Health Center. Blood samples were stored in the dark in a cool box. The bloods from both vacutainers were centrifuged (3000 g, 10 minutes) using portable centrifuges, serum/plasma was aliquoted and subsequently frozen at −20°C until transport (on dry ice) to the laboratory at NIN where they were stored at −80 °C until analysis.

Samples were analysed at VitMin Lab (Germany) for serum ferritin, transferrin receptor (sTfR), C-reactive protein (CRP), α-1-acid glycoprotein (AGP) and retinol binding protein (RBP) and at NIN laboratory for serum retinol and zinc. Serum ferritin, TfR, RBP, CRP and AGP concentrations were measured by VitMin Lab ELISA assays at VitMin Lab (Germany). Accuracy was tested by controls from CDC, USA and National Institute of Biological Standards and Control. Serum retinol concentration was determined by reverse-phase liquid chromatography with tandem mass spectrometry (Sciex Qtrap 6500+) with quality controls approved by CDC, US. Serum zinc concentration was analysed by a flame atomic absorption spectrophotometer (GBC, Avanta+) using trace element-free procedures, and powder free gloves (Latex Glove), and results were verified using reference materials (Liquicheck, Bio-Rad Laboratories, USA). The within-assay CV for serum ferritin, zinc, retinol, CRP and AGP ranged from 2.9 to 7.1 %, and between-assay variability was <10 % for all the biomarkers.

### Micronutrient deficiencies

Anaemia was defined as Hb concentrations <11.0 g/dL and <11.5 g/dL for children aged <5 and 5-9 years old, respectively (19). Different cut offs were applied to define iron deficiency (ID) in children of different age and inflammatory status. ID was defined as serum ferritin concentrations <12 µg/L (<5 years old) and <15 µg/L (5-9 years old) in children without inflammation. Whereas, cut offs of <30 µg/L (<5 years old) and <70 µg/L (5-9 years old) were applied for children with inflammation. Inflammation was defined as CRP >5 mg/L and/or AGP >1 g/L (20). Iron deficiency anaemia (IDA) was defined as the presence of both anaemia and ID. Low serum retinol was defined as serum retinol concentrations <0.70 µmol/L (21). Low serum zinc was defined as serum zinc concentration <9.9 μmol/L (morning, non-fasting) or <8.7 μmol/L (afternoon, non-fasting) according to International Zinc Nutrition Consultative Group (22). Total number of MNDs included anaemia, ID, low serum retinol and low serum zinc.

### Demographic and socioeconomic indicators

A structured questionnaire was used to collect demographic and socioeconomic data from the parents/caregivers including child’s age, sex, ethnicity (Kinh and other ethnicities), geographical areas (Northern mountains, Red River Delta, North Central and Central Coast, Central Highlands, Southeast, Mekong Delta River) and area of residence (urban, rural, and mountainous). Household wealth index was determined according to the Demographic and Health Survey (DHS) Wealth Index (23). It was computed based on household ownership of selected assets, materials used for housing construction, types of water access and sanitation facilities by the statisticians from NIN. The household wealth index was grouped into quintiles for further analysis: poorest (Q1), poorer (Q2), middle (Q3), richer (Q4) and richest (Q5) quintiles. The household wealth index was used as a proxy indicator for children’s socioeconomic status.

### Statistical analysis

All statistical analyses were conducted using STATA (version 18; Stata Corp LLC, Cary, NC, USA). All analyses were weighted and accounted for complex survey design using svyset commands to adjust for individual sampling weight, clustering and stratification. Baseline characteristics by area of residence were reported as mean ± SD for continuous variables and number (percentage) for categorical variables; they were compared using one-way between groups ANOVA with Bonferroni post hoc tests and Chi Squared test of association, respectively. Univariate logistic regressions were performed to determine the associations between demographic and socioeconomic factors and all forms of malnutrition. Both univariate and multivariate logistic regressions were performed to assess the associations between under- and over-nutrition and MNDs. Demographic and socioeconomic factors showed significant association in the univariate logistic regression analyses were included in the multivariate logistic regression analyses for adjustment. The covariates included age, sex, area of residence, wealth index and inflammation. Ethnicity and geographical area were not included due to multicollinearity. The crude odds ratio (OR) or adjusted OR (AOR) and respective 95% confidence intervals (CI) were reported. P value <0.05 was considered statistically significant.

## Result

General characteristics, anthropometric parameters, and micronutrients biomarkers of the study participants from rural, urban and mountainous areas are reported in **Table 1**. See **Table S1** for the number of participants with missing data for each variable. The average age ± SD of the study participants was 5 ± 3 years old and 48.5% were females. Nearly half of the children (52.4%) were aged 5-9 years old, whereas 31.6% and 16.0% of them were aged 2-4 and <2 years old, respectively. A significantly larger proportion of the children in urban areas were from the richest quintile (Q5) compared to those from the mountainous areas (17.3% vs. 1.6%; p= 0.005), and these children came mainly from the Kinh major ethnic group (93.5% vs. 36.3%; p<0.001). All anthropometric parameters except for HAZ were significantly higher in children from urban areas, compared to rural or/and mountainous areas (p<0.05). With respect to micronutrients biomarkers, concentrations of serum zinc were significantly higher, whereas serum transferrin receptor were significantly lower in children from urban areas (p<0.05). See **Table S2** for the mean (SD) age, anthropometric and biomarkers of micronutrients of the Vietnamese children by nutritional status.

**Table 1:**
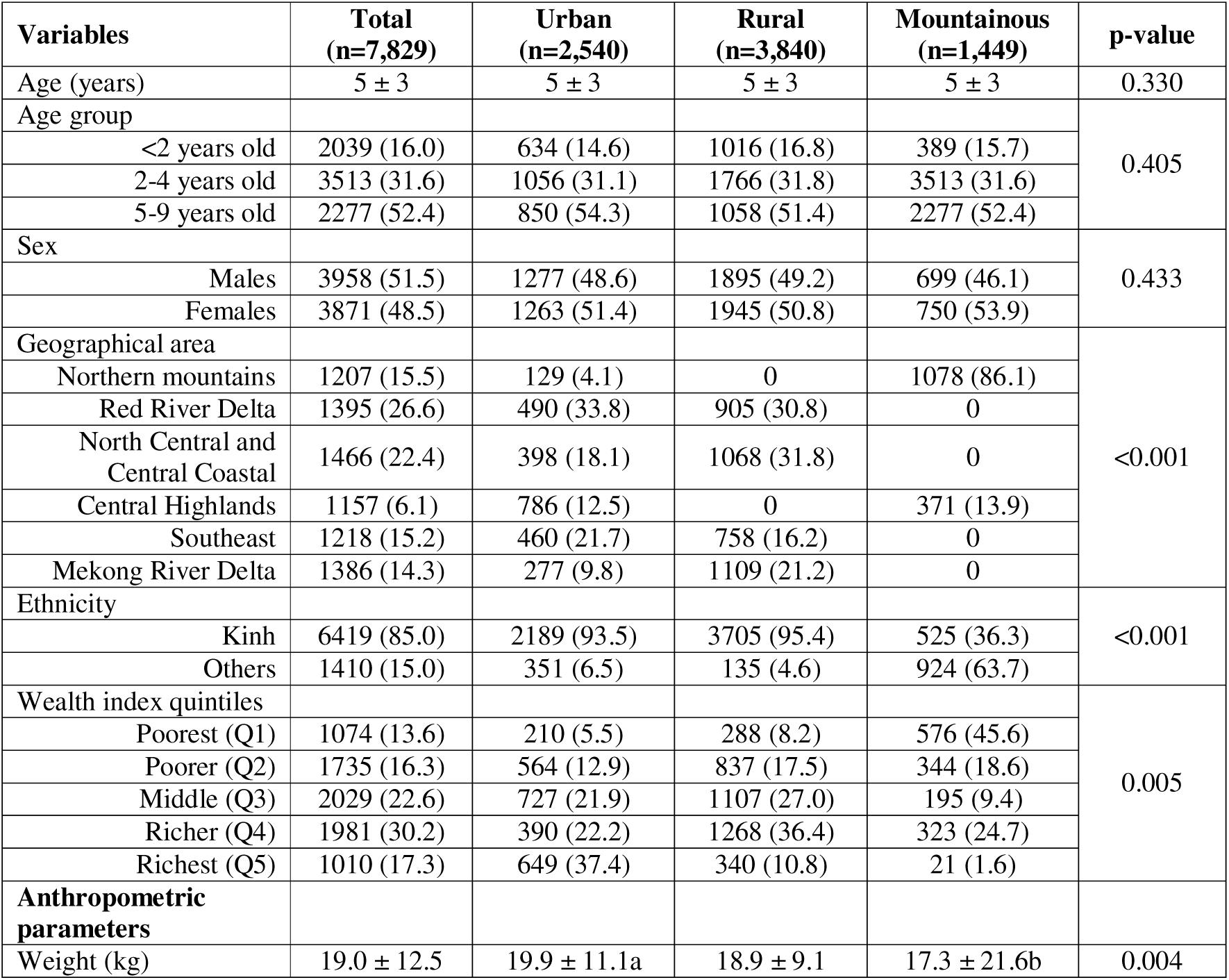
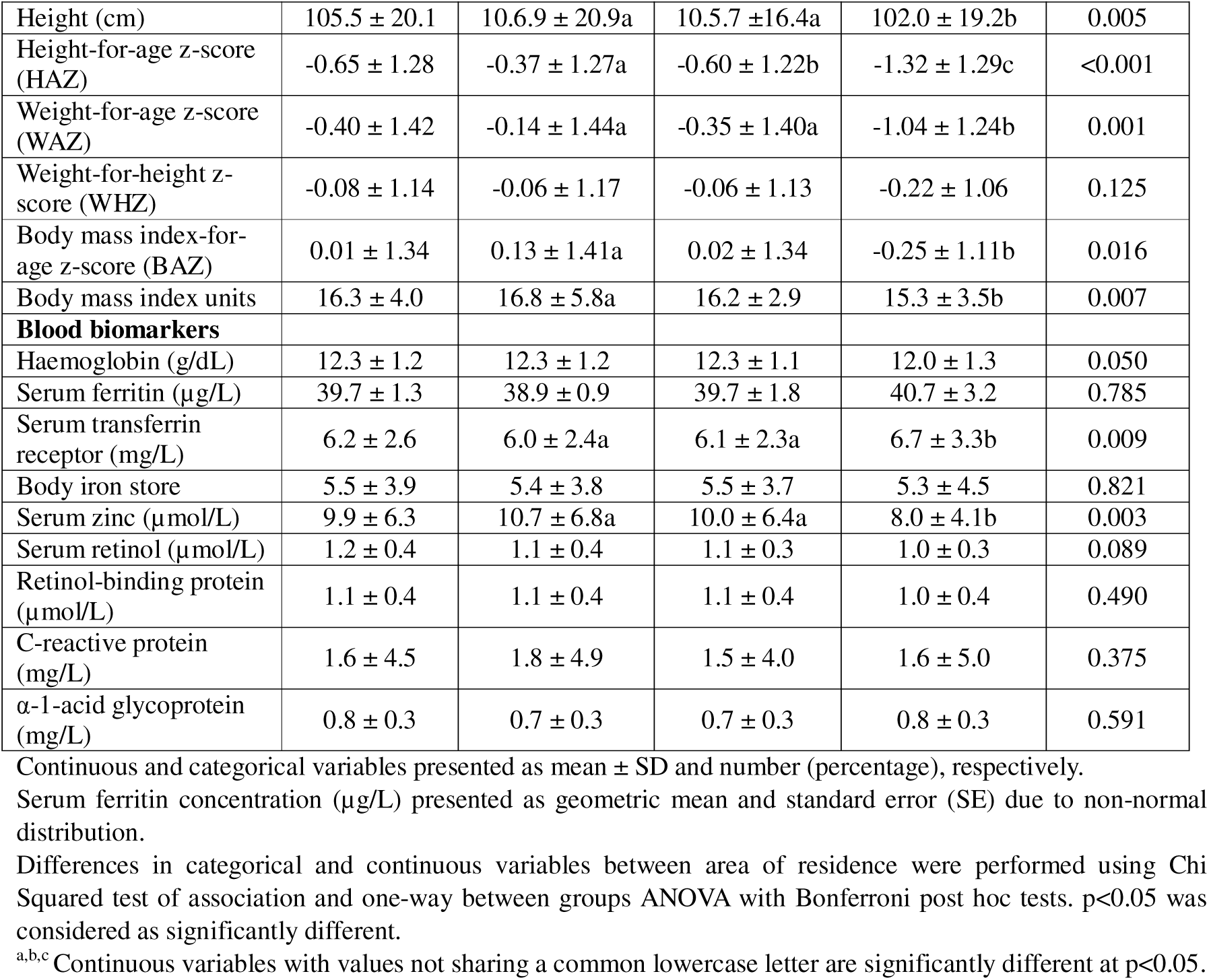
General characteristics, anthropometric parameters and micronutrients biomarkers of the Vietnamese children in this study, and by area of residence (n=7,829)

### Prevalence of undernutrition, overnutrition and MNDs by area of residence and age group

Overall, 12.7%, 10.5% and 4.7% of the children were either stunted, underweight or wasted/thin; while 7.3% and 7.1% were found overweight and obese, respectively. Low serum zinc was the most common MND investigated, affecting more than half (53.1%) of the Vietnamese children. Prevalence of ID, anaemia, IDA, and low serum retinol was 15.2%, 13.9, 5.8%, and 6.7% respectively. 16.2% of the children had high inflammatory status defined by CRP or/and AGP. When combining all four MNDs (anaemia, ID, low serum retinol and low serum zinc), 58.5% of the children had at least one of four MNDs (34.2% had one, 16.9% had two, 5.8% had three, and 1.6% had four MNDs). Prevalence of different forms of malnutrition by area of residence and age group are illustrated in **Figure 1**. Child stunting, underweight and wasting were more prevalent in mountainous areas compared to rural and urban areas. Prevalence of stunting was higher in children aged 2-4 years old (15% vs. 10.9%, and 11.0% in children aged <2 and 5-9 years old, respectively), particularly those from the mountainous areas (34.1%). Underweight and wasting were more prevalent among older children aged 5-9 years old (12.7% and 6.3%, respectively). Overweight and obesity were more common among older children aged 5-9 years old (23.0%) compared to those aged <2 (3.0%) and 2-4 (5.3%) years old, and was particularly evident in children aged 5-9 years old from the urban areas (31.4%).

**Figure 1:**
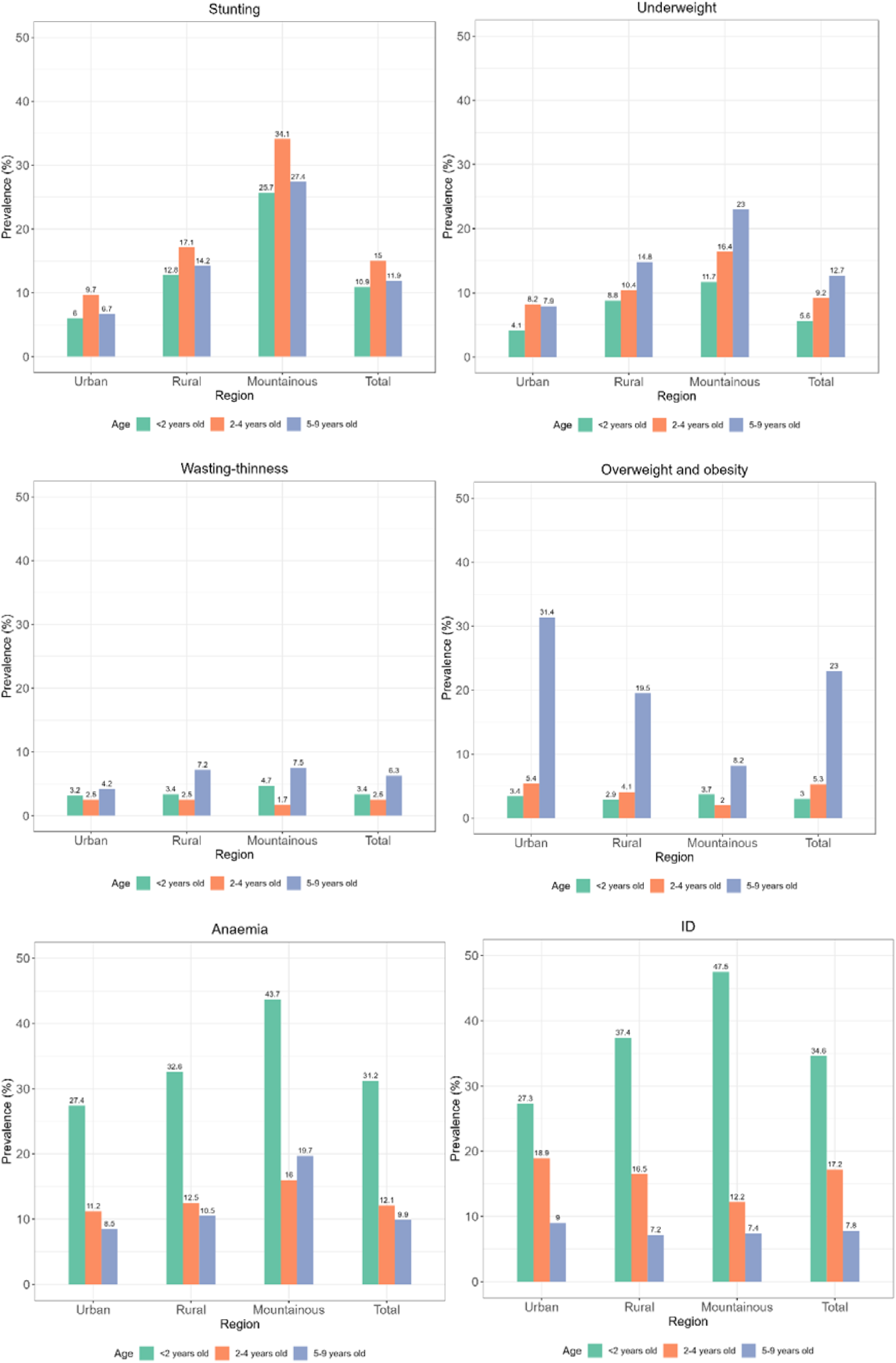
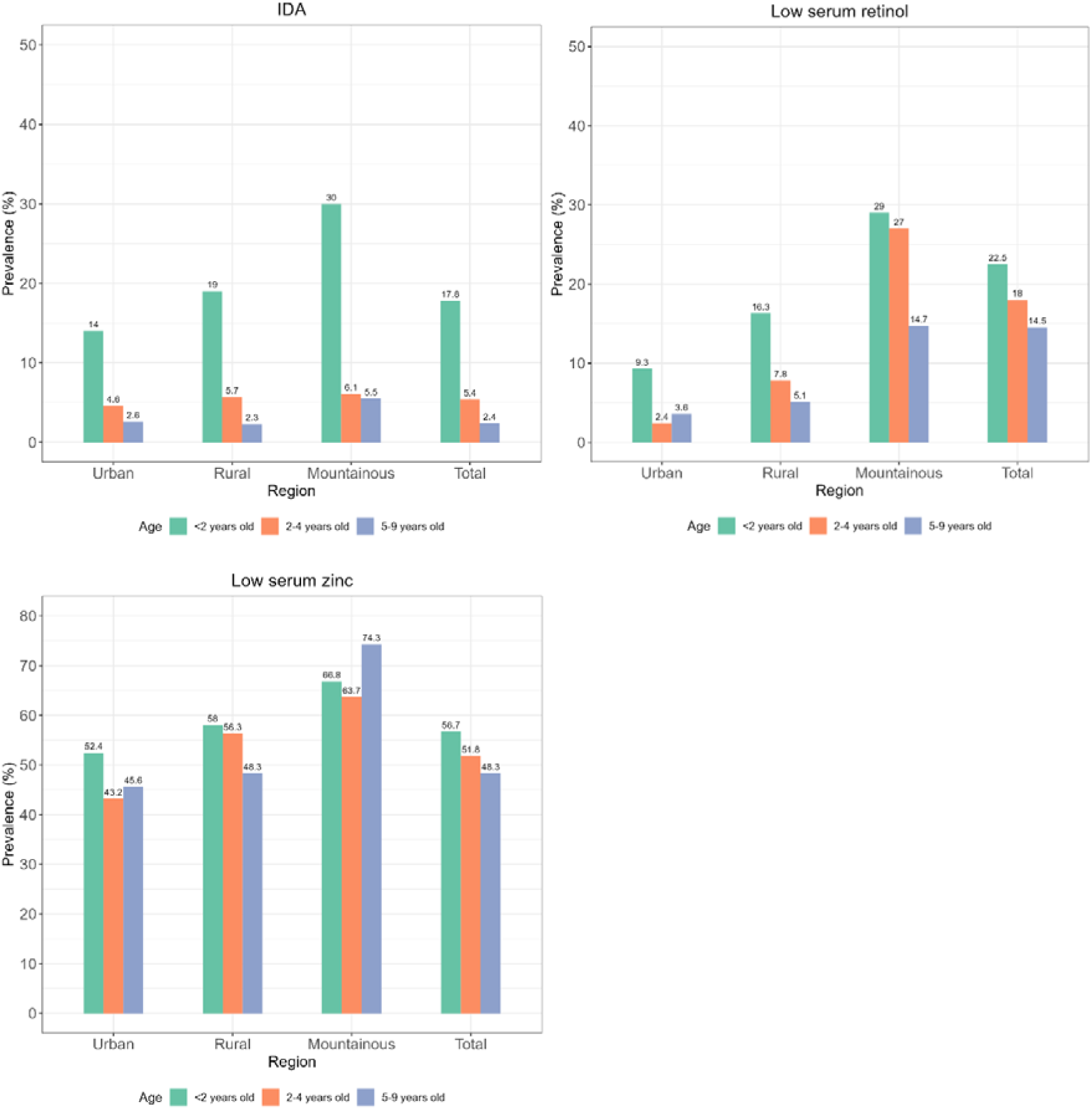
Prevalence of different forms of malnutrition by area of residence and age group among Vietnamese children. A) Stunting. B) Underweight. C) Wasting/thinness. D) Overweight and obesity. E) Anaemia. F) Iron deficiency. G) Iron deficiency anaemia. H) Low serum retinol. I) Low serum zinc. Prevalence was estimated based on sampling weight. ID, iron deficiency; IDA, iron deficiency anaemia.

Anaemia, ID, IDA, and low serum retinol were more prevalent among younger children aged <2 years old than in older children aged 2-4 and 5-9 years old, particularly if they were from the mountainous areas as opposed to the urban areas. Over two fifths of the children aged <2 years old from the mountainous areas were affected by anaemia (vs. 16.0% and 19.7% in children aged 2-4 and 5-9 years old), and ID (vs. 12.2% and 7.4% in children aged 2-4 and 5-9 years old), and about one third were affected by low serum retinol (vs. 22.7% and 14.7% in children aged 2-4 and 5-9 years old). Whereas higher prevalence of low serum zinc was observed in older children aged 5-9 years old, especially in children from the mountainous areas (74.3%) compared to the rural (48.3%) and urban (45.6%) areas.

### Associations of malnutrition and MNDs with demographic and socioeconomic indicators

The associations of demographic and socioeconomic indicators with malnutrition (under and overnutrition), and MNDs are reported in **Table 2** and **Table 3**, respectively. Compared to the youngest children (<2 years old), children aged 2-4 years old had a greater likelihood of being stunted [OR (95% CI): 1.43 (1.20, 1.72)] and school children (5-9 years old) had a greater likelihood of being wasted [2.48 (1.49, 4.14)] (**Table 2**). In addition, children aged 2-4 years old had significantly increased odds of underweight [1.70 (1.29, 2.23)] and overweight [1.75 (1.14, 2.70)], and the odds further increased in older children aged 5-9 years old [2.46 (1.70, 3.56); 10.02 (6.71, 14.97), respectively]. However, opposite trends were observed between age and MNDs. Compared to the youngest children, children aged 2-4 years old had significantly reduced odds of anaemia [0.30 (0.25, 0.37)], low serum retinol [0.32 (0.25, 0.42)] and low serum zinc [0.82 (0.69, 0.97)] (**Table 3**). The odds of anaemia [0.24 (0.19, 0.31)], ID [0.11 (0.07, 0.18)], IDA [0.11 (0.07, 0.18)] and low serum retinol [0.21 (0.15, 0.29)] were further reduced in older children aged 5-9 years old (**Table 3**).

**Table 2:** Univariate logistic regressions between malnutrition and demographic and socioeconomic indicators in Vietnamese children.

| Variables |  | Stunting<br>OR (95% CI) | Underweight<br>OR (95% CI) | Wasting/Thinness<br>OR (95% CI) | Overweight<br>OR (95% CI) |
| --- | --- | --- | --- | --- | --- |
| Age (<2 years) |  | Ref | Ref | Ref | Ref |
|  | 2-4 years | 1.43 (1.20, 1.72)*** | 1.70 (1.29, 2.23)** | 0.76 (0.49, 1.17) | 1.75 (1.14, 2.70)* |
|  | 5-9 years | 1.10 (0.84, 1.45) | 2.46 (1.70, 3.56)*** | 2.48 (1.49, 4.14)** | 10.02 (6.71, 14.97) *** |
| Sex (Females) |  | Ref | Ref | Ref | Ref |
|  | Males | 1.03 (0.85, 1.25) | 0.93 (0.76, 1.14) | 0.90 (0.61, 1.33) | 1.43 (1.16, 1.76)** |
| Area of residence (Urban) |  | Ref | Ref | Ref | Ref |
|  | Rural | 1.45 (0.86, 2.46) | 1.32 (0.86, 2.02) | 1.46 (0.90, 2.37) | 0.71 (0.46, 1.10) |
|  | Mountainous | 5.06 (1.81, 14.42)** | 2.98 (1.50, 5.92)** | 1.54 (0.59, 4.03) | 0.25 (0.08, 0.77)* |
| Ethnicity (Kinh) |  | Ref | Ref | Ref | Ref |
|  | Others | 5.94 (3.78, 9.36)*** | 3.68 (2.81, 4.82)*** | 1.56 (0.71, 3.44) | 0.15 (0.07, 0.34)*** |
| Geographical area (Northern mountains) |  | Ref | Ref | Ref | Ref |
|  | Red River Delta | 0.29 (0.09, 0.96)* | 0.44 (0.19, 0.99)* | 1.01 (0.52, 1.96) | 1.87 (0.97, 3.60) |
|  | North Central and Central Coastal | 0.50 (0.14, 1.75) | 0.55 (0.21, 1.45) | 0.86 (0.48, 1.56) | 1.62 (0.86, 3.07) |
|  | Central Highlands | 0.76 (0.17, 3.42) | 1.09 (0.35, 3.36) | 2.03 (0.74, 5.57) | 1.38 (0.54, 3.50) |
|  | Southeast | 0.19 (0.05, 0.74)* | 0.21 (0.09, 0.47)** | 0.71 (0.32, 1.60) | 3.88 (2.07, 7.25)*** |
|  | Mekong River Delta | 0.28 (0.09, 0.91)* | 0.65 (0.29, 1.49) | 1.25 (0.75, 2.10) | 2.90 (1.46, 5.79)** |
| Wealth index quintiles (Poorest; Q1) |  | Ref | Ref | Ref | Ref |
|  | Poorer (Q2) | 0.40 (0.19, 0.82)* | 0.46 (0.31, 0.68)** | 1.02 (0.38, 2.71) | 2.94 (1.20, 7.21)* |
|  | Middle (Q3) | 0.23 (0.11, 0.50)** | 0.27 (0.19, 0.40)*** | 0.59 (0.23, 1.49) | 2.56 (1.04, 6.29)* |
|  | Richer (Q4) | 0.20 (0.09, 0.43)*** | 0.26 (0.16, 0.43)*** | 0.65 (0.25, 1.67) | 2.65 (1.13, 6.21)* |
|  | Richest (Q5) | 0.13 (0.05, 0.32)*** | 0.17 (0.10, 0.29)*** | 0.50 (0.20, 1.27) | 2.91 (1.20, 7.05)* |

CI, confidence interval; OR, odds ratio; ref, reference.
Odds ratio and 95% confidence interval were estimated using univariate logistic regression analyses. 'Non-malnourished' groups were used as the reference group.
\* p<0.05; \*\* p<0.01; \*\*\* p<0.001.

**Table 3:**
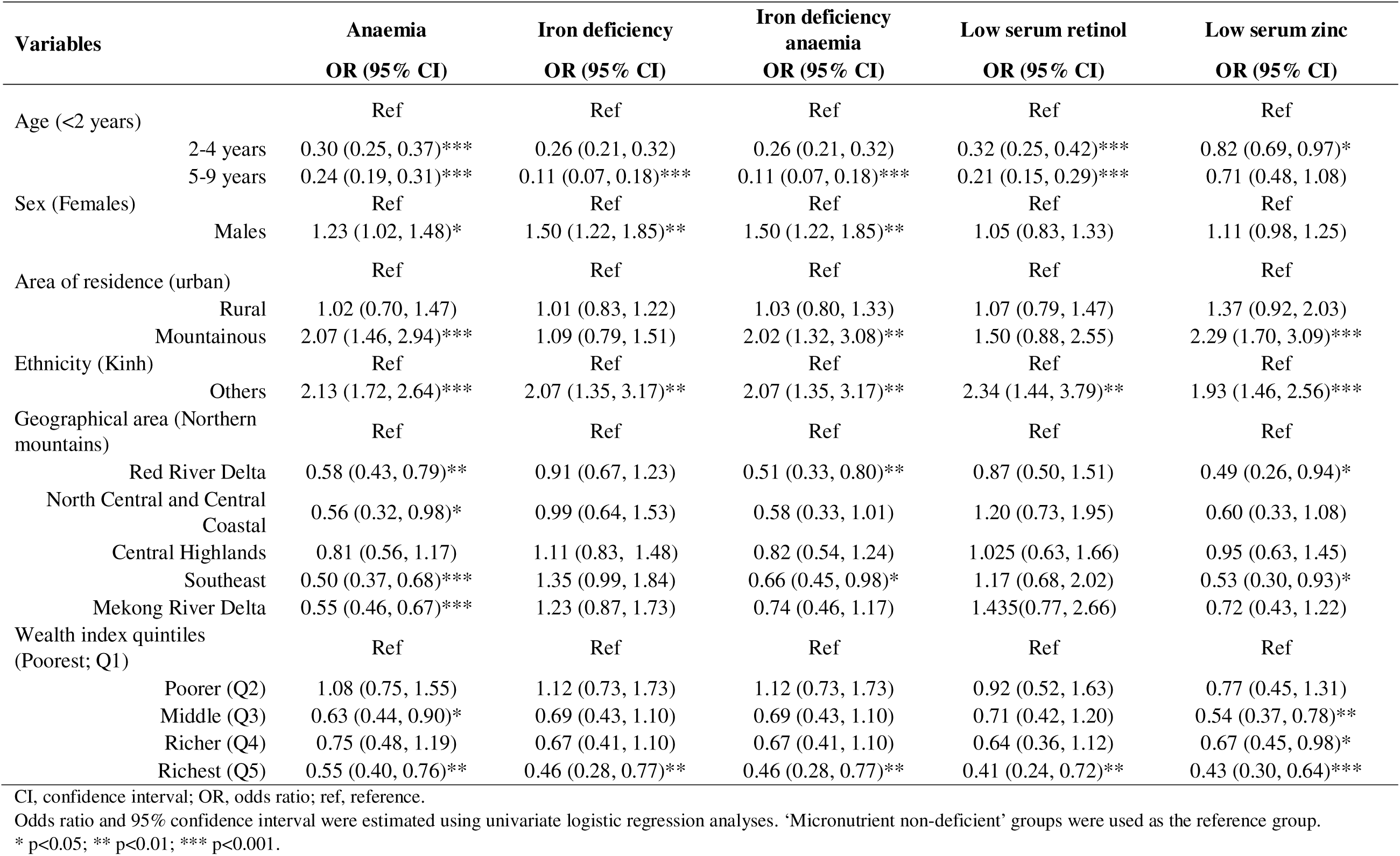
Univariate logistic regressions between micronutrient deficiencies and demographic and socioeconomic indicators in Vietnamese children.

Males were more likely to be overweight [1.43 (1.16, 1.76)] and affected by anaemia [1.23 (1.02, 1.48)], ID [1.50 (1.22, 1.85)] and IDA [1.50 (1.22, 1.85)] compared to females (**Table 2 and 3**). Children from the mountainous areas had increased odds of stunting [5.06 (1.81, 14.42)], underweight [2.98 (1.50, 5.92)], low serum zinc [2.29 (1.70, 3.09)] and anaemia [2.07 (1.46, 2.94)] compared to the urban areas. Children from the ethnic minorities had increased odds of stunting [5.94 (3.78, 9.36)], underweight [3.68 (2.81, 4.82)] and all MNDs investigated (OR ranging from 1.93-2.34), and reduced odds of overweight [0.15 (0.07, 0.34)]. The odds of being stunted and underweight were significantly decreased across the wealth index quintiles in a dose-dependent manner, with children from the richest quintile (Q5) were most protected against stunting [0.13 (0.05, 0.32)], and underweight [0.17 (0.10, 0.29)], compared to the poorest quintile (Q1) (**Table 2**). In contrast, children from greater wealth index quintiles (Q2-Q5) had significantly increased odds of overweight (OR ranging from 2.6-2.9). In addition, children from the richest quintile (Q5) were also protected against anaemia [0.55 (0.40, 0.76)], ID [0.46 (0.28, 0.77)], and IDA [0.46 (0.28, 0.77)], low serum retinol [0.41 (0.24, 0.72)] and low serum zinc [0.43 (0.30, 0.64)] (**Table 3**).

Compared to Northern mountains, children from Southeast and Mekong River Delta had reduced odds of stunting [0.19 (0.05, 0.74); and 0.28 (0.09, 0.91)], but had increased odds of overweight [3.88 (2.07, 7.25); and 2.90 (1.46, 5.79)], respectively (**Table 2**). Children from Red River Delta and Southeast had also reduced odds of underweight [0.44 (0.19, 0.99); and 0.21 (0.09, 0.47), respectively]. Moreover, children from the Red River Delta and Southeast were less likely to be affected by anaemia, IDA and low serum zinc (OR ranging from 0.49-0.66) (**Table 3**).

### Associations of undernutrition and overnutrition with micronutrient deficiencies

In the adjusted multivariate logistic regressions, children with anaemia [AOR (95% CI): 1.43 (1.13, 1.82)], IDA [1.53 (1.22, 1.93)] and low serum zinc [1.44 (1.11, 1.88)] had significantly increased odds of stunting compared to the non-deficient children (**Table 4**). Children with low serum retinol [1.71 (1.21, 2.42)] and low serum zinc [1.51 (1.13, 2.01)] had increased odds of being underweight, and children with low serum retinol also had reduced odds of being overweight [0.43 (0.27, 0.68)]. When combining the number of MNDs occurring in each child, we found that children with three MNDs [1.96 (1.14, 3.37)] and children with four MNDs [1.84 (1.08, 3.13)] had increased odds of stunting, compared to those without MNDs. Similarly, children with two, three and four MNDs had increased odds of being underweight in a dose-dependent manner [1.48 (1.01, 2.17); 1.81 (1.17, 2.81); and 2.26 (1.17, 4.35), respectively]. Children with one, two and three MNDs were less likely to be overweight or obese [0.60 (0.44, 0.81); 0.38 (0.23, 0.63); and 0.44 (0.23, 0.85), respectively].

**Table 4:** Univariate and multivariate logistic regressions between malnutrition and micronutrient deficiencies in Vietnamese children.

| Variables | Stunting |  | Underweight |  | Wasting/Thinness |  | Overweight/obese |  |
| --- | --- | --- | --- | --- | --- | --- | --- | --- |
|  | OR (95% CI) | AOR (95% CI) | OR (95% CI) | AOR (95% CI) | OR (95% CI) | AOR (95% CI) | OR (95% CI) | AOR (95% CI) |
| Anaemia (No)<sup>b</sup> | Ref | Ref | Ref | Ref | Ref | Ref | Ref | Ref |
| Yes | 1.39 (1.14, 1.70)** | 1.43 (1.13, 1.82)** | 1.09 (0.84, 1.42) | 1.33 (0.99, 1.78) | 0.79 (0.51, 1.24) | 0.88 (0.53, 1.47) | 0.57 (0.38, 0.85)** | 0.76 (0.50, 1.15) |
| Iron deficiency (No)<sup>a</sup> | Ref | Ref | Ref | Ref | Ref | Ref | Ref | Ref |
| Yes | 1.17 (0.90, 1.52) | 1.11 (0.88, 1.40) | 0.81 (0.52, 1.25) | 1.01 (0.68, 1.51) | 0.49 (0.25, 0.95)* | 0.65 (0.33, 1.26) | 0.63 (0.50, 0.79)** | 1.07 (0.79, 1.44) |
| Iron deficiency anaemia (No)<sup>a</sup> | Ref | Ref | Ref | Ref | Ref | Ref | Ref | Ref |
| Yes | 1.52 (1.05, 2.18)* | 1.53 (1.22, 1.93)* | 1.00 (0.60, 1.67) | 1.33 (0.76, 2.24) | 0.44 (0.25, 0.76)** | 0.56 (0.32, 1.01) | 0.54 (0.35, 0.85)* | 1.03 (0.58, 1.84) |
| Low serum retinol (No)<sup>b</sup> | Ref | Ref | Ref | Ref | Ref | Ref | Ref | Ref |
| Yes | 1.55 (1.02, 2.36)* | 1.40 (0.86, 2.30) | 1.47 (1.04, 2.07)* | 1.71 (1.21, 2.42)** | 1.02 (0.60, 1.72) | 1.21 (0.73, 1.99) | 0.34 (0.20, 0.57)*** | 0.43 (0.27, 0.68)*** |
| Low serum zinc (No)<sup>b</sup> | Ref | Ref | Ref | Ref | Ref | Ref | Ref | Ref |
| Yes | 1.49 (1.12, 1.97)** | 1.44 (1.11, 1.88)** | 1.43 (1.07, 1.91)* | 1.51 (1.13, 2.01)** | 1.15 (0.71, 1.87) | 1.08 (0.66, 1.76) | 0.63 (0.37, 1.05) | 0.65 (0.38, 1.10) |
| Total number of MND (None)<sup>b</sup> | Ref | Ref | Ref | Ref | Ref | Ref | Ref | Ref |
| 1 | 1.02 (0.74, 1.41) | 1.05 (0.72, 1.51) | 0.88 (0.68, 1.15) | 0.98 (0.72, 1.34) | 0.71 (0.42, 1.20) | 0.87 (0.53, 1.44) | 0.48 (0.36, 0.63)*** | 0.60 (0.44, 0.81)** |
| 2 | 1.51 (1.07, 2.14)* | 1.58 (0.99, 2.52) | 1.05 (0.76, 1.46) | 1.48 (1.01, 2.17)* | 0.69 (0.40, 1.21) | 1.01 (0.57, 1.78) | 0.22 (0.15, 0.31)*** | 0.38 (0.23, 0.63)** |
| 3 | 1.67 (1.17, 2.40)** | 1.96 (1.14, 3.37)* | 0.95 (0.58, 1.54) | 1.81 (1.17, 2.81)* | 0.59 (0.29, 1.22) | 0.92 (0.48, 1.77) | 0.16 (0.09, 0.29)*** | 0.44 (0.23, 0.85)* |
| 4 | 1.58 (1.04, 2.40)* | 1.84 (1.08, 3.13)* | 1.05 (0.59, 1.89) | 2.26 (1.17, 4.35)* | 0.30 (0.07, 1.34) | 0.49 (0.12, 2.06) | 0.31 (0.12, 0.79)* | 1.12 (0.43, 2.92) |

AOR, adjusted odds ratio; CI, confidence interval; MND, micronutrient deficiencies; OR, odds ratio; ref, reference.
Univariate and multivariate logistic regression analyses were performed and reported as OR (95% CI) and AOR (95% CI), respectively. 'Micronutrient non-deficient' groups were used as the
reference group.
<sup>a</sup>adjusted by age, sex and area of residence; and <sup>b</sup>adjusted by age, sex, area of residence and inflammation.
\* p<0.05; \*\* p<0.01; \*\*\* p<0.001.

## Discussion

### Prevalence of undernutrition, overnutrition, and MNDs

To the best of our knowledge, this is the first reported population-based study in Vietnam to assess MNDs (anaemia, iron, vitamin A and zinc deficiencies) in children from a nationally representative nutrition survey that also considers the associations with nutritional status (defined by undernutrition and overnutrition). Findings from the GNS 2020 indicate that the TBM remains a major public health issue for Vietnamese children. This survey shows that the prevalence of stunting in children aged <2, and 2-4 years old was 10.9% and 15.0%, respectively, in 2020, suggesting that Vietnam is now on course to achieve Global Nutrition Targets which is to reduce the prevalence of stunting in children aged <5 years old by 40% in 2025 compared to 2012 (<20%) (6, 24). Child stunting is now considered as ‘medium’ public health significance (<20%) in Vietnam according to WHO (24). In contrast, Vietnam is off course to meet the Global Nutrition Targets 2025 for childhood overweight and obesity, which is to prevent the figure from increasing. Increasing overweight and obesity in Vietnam is evident, particularly in school-aged children of urban areas (31%). This prevalence has been substantially increased compared to 8.5% in 2010 in children of same age (9). MNDs (low serum zinc, anaemia and ID) were widespread among children in Vietnam, although low serum retinol was of less concern (<7%). In line with previous findings, this survey reveals that more than half of the Vietnamese children aged 6 months to 9 years old had low serum zinc status (25, 26). Anaemia and ID affected 15% and 13% of the study participants, respectively.

### Demographic variation and socioeconomic inequalities in undernutrition, overnutrition and MNDs

Our findings demonstrated regional and demographic variation and socioeconomic inequalities in child malnutrition in Vietnam. Prevalence of childhood overweight and obesity was higher in males, living in urban areas and richer families. This may be due to better economic growth, and improved standard of living and nutrition (27). Evidence from reviews and longitudinal studies reported that unhealthy diets especially consumption of sugary sweetened beverages and fast foods, physical inactive, and lack of sleep are the major risk factors for overweight and obesity in Vietnamese school-aged children and adolescents, particularly in developed areas (9, 28, 29). In addition, maternal obesity, gestational diabetes during pregnancy, and suboptimal feeding practices are significant contributors for early childhood overweight (9, 28, 29). However, there is limited data on the key determinants and effective strategies to address childhood overweight and obesity in this region, thus future research is warranted.

In this study, the prevalence of stunting, underweight and MNDs was high in infants and young children, particularly in children from the ethnic minorities, rural and mountainous areas, and poorer families. Previous evidence reported that ethnic minorities living in poor and remote rural and mountainous areas had limited economic, social, or cultural support including education and health services compared to the major ethnic groups (Kinh or Hoa) (10, 30). This results in large disparities in nutrition outcomes, and persistent high prevalence of child undernutrition and MNDs (31). The Food Agriculture Organization (FAO) Report also postulated that potential explanatory factors include climate change, conflict and slowed economic development, all of which contributes to an increased socioeconomic inequality in food insecurity and malnutrition between urban and rural regions (32). More efforts and attention are needed to reduce inequality gap in relation to socioeconomic status among ethnic minorities or vulnerable populations living in the poor, remote mountainous areas (33). Thus, national policies and intervention programmes should address age-specific, sex-specific, geographical and socioeconomic disparities to ensure it reaches people at greatest risk and in greatest need.

### Low serum zinc

More than half of the Vietnamese children were affected by low serum zinc status. However, caution needs to be taken when assessing and interpreting zinc deficiency using plasma or serum samples, as many factors may affect plasma or serum zinc concentration such as the time when blood was collected, meal consumption, fasting or non-fasting status and inflammation (34). Another concern is that the potential contamination of blood samples may underestimate the prevalence of zinc deficiency. To date, there is no consensus on which indicators are best to assess zinc status, making it difficult to compare and assess the extent of zinc deficiency of a population (34). Nonetheless, plasma or serum zinc is still a frequently used biomarker at population level (35). Household Living Standards Survey 2016 reported that rice was the most common food item consumed by Vietnamese population (almost 300g per person per day), and it was the only staple food consumed by nearly all households (36). However, rice contains very low amount of zinc and high amount of phytic acid, which will bind to zinc and form phytate salts that further inhibit zinc and iron absorption (37). Previous evidence suggested that animal-based foods e.g., fish, poultry, red meat or liver and dairy products e.g., milk and cheese are key contributors to zinc intake in this population (37, 38), thus increase consumption of these food items may have positive impact on zinc status. More effective public health solutions can be achieved through zinc supplementations (39) or bio-fortification of zinc rich foods e.g., zinc bio-fortified maize, wheat or rice (40).

### Low serum retinol

Low serum retinol can lead to impaired immune system which subsequently increases the risk of infectious diseases, and child morbidity and mortality (41). In 2012, Laillou et al. reported that 11.8% and 11.9% of the children aged <2 and 2-5 years old had low serum retinol (26). Concerningly, within a decade, the prevalence of low serum retinol has increased substantially, with 22.5% children aged <2 years old, and 18.0% of children aged 2-5 years old in this study were found to have low serum retinol, respectively. However, such discrepancies may be partly due to urban-rural difference as larger proportion of children in this survey were from rural areas (77.6% vs. 48.4%). Notably, Vietnam has stipulated mandatory vitamin A fortification in sugar and vegetable oil (42) and implemented vitamin A supplementation programme targeted at children aged 6-36 months since 1997 with a population coverage of 99% in 2000, which may have substantially contributed to the reduction of vitamin A deficiency (43). However, the coverage of vitamin A distribution in children from poor, remote and mountainous areas is still lower than other areas (43). This may explain the high prevalence of low serum retinol in mountainous areas (14.7-29.0%) than in urban areas (<10%). It is therefore relevant that findings from this study inform the current design of vitamin A supplementation or fortification programmes, to ensure it reaches the vulnerable populations that are most in need. Promotion of the consumption of vitamin A-rich fruits and vegetables should also be encouraged to sustainably combat vitamin A deficiency. Integration of deworming treatments into an existing vitamin A supplementation may be considered to improve child’s vitamin A status especially in areas with high burden of infection.

### Anaemia and iron deficiency (ID)

Our findings underscore that anaemia and ID were more prevalent among children aged <2 years old, males and those living in the mountainous areas. Higher prevalence of anaemia in children aged <2 years old have been reported in many other populations (44, 45), but not all (46). In the first 4–6 months of life, iron supply acquired in utero is sufficient to meet requirements of infants with unimpaired intrauterine supply, whereas after 6 months, the intrauterine-supplied iron becomes insufficient due to accelerated growth and development, leading to increased risk of deficiency (47). Due to immaturity of gut microbiome and immune system, infants and young children are more susceptible to infections and diseases, leading to increased risk of nutritional deficiencies (48). Increased risk of anemia in males during early childhood than in females have also been reported in many studies (49–52). Previous studies reported significantly lower cord serum ferritin levels and higher cord sTfR in males than in females at birth and at first 2 years of life, which suggests that males were at greater risk of low iron status (53). Although the underlying mechanisms are unknown, but this mechanism may be explained by hormone-mediated differences in metabolism e.g., increased erythropoietin activity for red blood cell production in males may result in low iron store (54). Differences in the proportion of lean and fat mass and weight gain (e.g., greater weight gain in males) may also indirectly affect iron metabolism (55).

ID is frequently reported as the main nutritional contributor to anaemia, but the aetiology of anemia is complex, multifactorial and context-specific (56). Our findings indicate that nearly one third of the anaemia cases in Vietnamese children were attributable to ID. The proportion of anaemia attributable to ID was highest in children aged <2 years old (57%) compared to older children aged 5-9 years old (24.4%). This finding was in line with previous evidence reported that the prevalence of anaemia attributable to ID was highest in the 1–4 age group, and then gradually decreased up to the 25–29 age group (50). Cautious need to be taken when interpretating the results as unmeasured factors such as high burden of disease and infection or haemoglobinopathies may overestimate the proportion of anaemia accounted for by ID (57). Other important known risk factors of child anaemia include nutritional deficiencies (e.g., iron, vitamin A, folate, vitamin B12), environmental factors, chronic comorbidities (e.g., infection, intestinal parasites, malaria), and congenital or genetic disorders (e.g., sickle cell, α-thalassemia), which are further complicated by socioeconomic and ecological risk factors (58). Therefore, these risk factors should be considered when investigating the underlying causes of anaemia, to develop appropriate and effective integrated interventions to prevent and treat child anemia in this population.

### Associations between MNDs and undernutrition and overnutrition

Our findings showed that MNDs were highly associated with child stunting and underweight. Inadequate intake of micronutrients especially iron, zinc and retinol, A and C may result in growth retardation (59). Poor energy and nutrient diets are known as the main cause of both stunting and MNDs, other factors include those associated with poverty or poor socioeconomic development such as poor sanitation and hygiene, infection, poor maternal nutrition and feeding practices (60). Despite excessive energy consumption, previous evidence demonstrated that children living with overweight and obesity were more likely to be affected by MNDs, and this may be primarily due to poor nutrient energy-dense diets that low in micronutrient content (61). In contrast, opposite results were found in our study, overweight was protective against the development of multiple forms of MNDs. The possible explanations include overweight and obese children consume more to meet their nutritional requirements or the fortification of certain processed foods (62). To date, the direction of association between overnutrition and MNDs are still unknown (63), it is unclear whether specific MNDs contribute to the increasing risk of greater obesity and adiposity, or increased overweight and adiposity resulted in specific MNDs, or maybe both. The link between overnutrition and MNDs is further complicated by the nutrient density as diets containing similar energy content can vary in terms of micronutrients that they provide (64). Thus, future research is warranted to shed light the relationship between overnutrition and MNDs.

### Strategies to address child undernutrition, MNDs, and overnutrition

Child undernutrition and MNDs were more prevalent among younger children aged <2 years old in this study, highlighting the importance of optimising nutrition in early life in alleviating undernutrition and MNDs for Vietnamese children. Adequate maternal nutrition and early childhood nutrition especially during the first 1000 days of life, is essential to ensure optimal growth and development of children to their full potential (65). Thus, interventions targeted at the first 1000 days of life such as multi-micronutrients supplementation and food fortification during pregnancy and infancy, nutrition education to optimise infant and young children feeding (IYCF) practices (e.g., breastfeeding and complementary feeding) and promoting dietary diversification using locally available micronutrient-rich foods may help to tackle multiple forms of malnutrition in infant and young children (66).

In recent years the Vietnamese government has published and implemented a National Nutrition Strategy 2021-2030 with a vision to 2040, which included targets and indicators on overweight and obesity in children and adolescents (9). However, the current policies and guidelines in Vietnam were not specifically designed to address childhood overweight and obesity, rather all the programmes were integrated or designed as a small part of broader public health strategies (67). Such policies and guidelines included tax for sugary-sweetened beverages (SBB), nutrition labelling, restriction on unhealthy food marketing in school campus, physical activity programmes and national dietary guidelines. More obesity prevention research is warranted to provide sufficient evidence to inform policy decision to address childhood overweight and obesity. It is also essential to raise public awareness about the consequences and risk factors of overweight and obesity to generate demands for healthier options and strengthen nutrition education to promote healthy eating behaviour and lifestyle in children and parents/caregivers.

### Strengths and limitations

This study had some notable strengths. The nationally representative sample of Vietnamese children allows the findings to be generalized across the country. Stratified analyses by age groups, area of residence and sex provide insights on the vulnerable population in Vietnam who are at the greatest risk of developing multiple forms of malnutrition. This finding helps to better tailor interventions, ensuring they reach the populations that most in needs to improve outcomes. There were few limitations in this study. As in all observational studies, this study was limited by confounding and causality should not be inferred. Food consumption data in children was not available in the GNS 2020, thus analysis of the association between food intake and child nutritional status was not possible.

## Conclusion

Our findings indicate that the TBM is becoming a major public health concern for Vietnamese children. Demographic variation and socioeconomic inequalities in TBM remain a critical challenge for policymakers. Child undernutrition and MNDs are more prevalent in infants and young children, particularly in children from the ethnic minorities, rural and mountainous areas and poor families. Whereas increasing childhood overweight and obesity are evident, particularly in males, school aged children from urban areas and rich families. Thus, national policies and intervention programmes in Vietnam should address age-specific, sex-specific, geographical and socioeconomic disparities to ensure equitable access and opportunities for all, and to accelerate progress in improving child nutrition. Existing food and nutrition programmes and policies need to be redesigned to use a multisectoral, double-duty approach to simultaneously address all forms of child malnutrition.

## Data availability statement

The datasets presented in this article are not publicly available. This dataset is the property of the National Institute of Nutrition (NIN), Vietnam, and is available for research purposes on reasonable request. Requests to access the dataset should be directed to.

## Ethics statement

Ethical approval was not required for secondary analysis in this study. The GNS 2020 was reviewed and approved by the Ethical Committee of the National Institute of Nutrition (NIN), Ministry of Health, Vietnam. All methods were conducted in accordance with the guidelines laid down in the Declaration of Helsinki. Written informed consent was obtained from each participant prior to data collection by NIN. Research team from the University of Leeds (UoL) (PYT, VSS, and XT) were permitted to access a sub-set of the anonymised GNS 2020 dataset. The UoL research team created the do files with STATA commands for analysis, and SN and TDT from the NIN were responsible for checking and running the commands on the full GNS 2020 dataset in STATA.

## Acknowledgment

This work was conducted under a partnership agreement between the University of Leeds, National Institute of Nutrition, Vietnam, and International Life Sciences Institute Southeast Asian Region (ILSI SEAR). The authors would also like to extend their gratitude to the National Institute of Nutrition, Ministry of Health, Vietnam, for their support in providing access to the GNS 2020 dataset.

## Contributors

YYG, TNT, VKT, LD and PYT contributed to developing the concept of this work and the secondary analysis plan using data from GNS 2020. NTT and VTK contributed to the methodology design and local implementation of the study. DTT and NDS prepared the GNS 2020 dataset required for analysis. PYT, VSS, XT and NDS contributed to data cleaning and checking, performed statistical analysis, and interpreted and revised results. XT contributed to data visualisation (graph). PYT and VSS drafted the first version of manuscript. All authors provided input into the interpretation of the analyses presented in the manuscript. All authors edited and approved the final manuscript. YYG had oversight of this work. YYG (Principal investigator) and LD, JBM, SC, HE and XL (co-investigators) acquired funding for this work.

## Patient and public involvement

Patients and/or the public were not involved in the design, or conduct, or reporting, or dissemination plans of this research.

