## Supplementary Table 1 and 2 for "Demographic variation and socioeconomic inequalities associated with the triple burden of malnutrition in Vietnamese children aged 6 months to 9 years old: Findings from the Vietnamese General Nutrition Survey 2020"

**Supplementary materials**

**Supplementary Table 1: Number of observations and missing data for each variable**

| **Variable** | **Number of observations** | **Missing data (%)** |
| --- | --- | --- |
| **Demographic and socioeconomic indicators** |  |  |
| Age | 7,829 | 0 |
| Sex | 7,829 | 0 |
| Area of residence | 7,829 | 0 |
| Geographical area | 7,829 | 0 |
| Ethnicity | 7,829 | 0 |
| Wealth index | 7,829 | 0 |
| **Nutritional status** |  |  |
| Stunting | 6,796 | 13.2 |
| Underweight | 6,813 | 13.0 |
| Wasting/thinness | 6,790 | 13.3 |
| Overweight | 6,790 | 13.3 |
| **Micronutrient deficiencies** |  |  |
| Anaemia | 7,249 | 7.4 |
| Iron deficiency | 6,800 | 13.1 |
| Iron deficiency anaemia | 7,249 | 7.4 |
| Low serum retinol | 7,124 | 9.0 |
| Low serum zinc | 5,271 | 32.7 |
| Inflammation (CRP and AGP) | 6,831 | 12.7 |

AGP, α1-acid glycoprotein; CRP, c-reactive protein.

Supplementary Table 2: General characteristics, anthropometric parameters, and micronutrient biomarkers of the Vietnamese children by nutritional status

| **Variables** | **Total** | **Non- stunted** | **Stunted** | **p value** | **Non- underweight** | **Underweight** | **p value** | **Non- wasted** | **Wasted** | **p value** | **Non- overweight** | **Overweight** | **p value** |
| --- | --- | --- | --- | --- | --- | --- | --- | --- | --- | --- | --- | --- | --- |
| **Demographic and socioeconomic indicators** | | | | | | | | | | | | | |
| Age (years) | 5 ± 3 | 5 ± 3 | 5 ± 3 |  | 5 ± 3 | 6 ± 3 | *** | 5 ± 3 | 6 ± 3 | ** | 5 ± 3 | 7 ± 2 | *** |
| Age group |  |  |  |  |  |  |  |  |  |  |  |  |  |
| <2 years | 2039 (16.0) | 1521 (15.8) | 213 (13.3) | ** | 1622 (16.3) | 117 (8.2) | *** | 1669 (15.6) | 64 (11.1) | *** | 1681 (17.4) | 52 (3.2) | *** |
| 2-4 years | 3513 (31.6) | 2602 (30.4) | 446 (36.7) |  | 2783 (31.6) | 272 (27.1) |  | 2951 (31.7) | 83 (16.6) |  | 2855 (34.3) | 179 (11.3) |  |
| 5-9 years | 2277 (52.4) | 1794 (53.8) | 220 (50.0) |  | 1766 (52.1) | 253 (64.7) |  | 1895 (52.7) | 128 (72.3) |  | 1565 (48.2) | 458 (85.5) |  |
| Sex |  |  |  |  |  |  |  |  |  |  |  |  |  |
| Males | 3958 (51.5) | 2935 (48.3) | 423 (47.5) |  | 3048 (47.8) | 315 (49.6) |  | 3220 (48) | 142 (52.1) |  | 3064 (49.5) | 298 (40.5) | ** |
| Females | 3871 (48.5) | 2982 (51.7) | 456 (52.5) |  | 3123 (52.2) | 327 (50.4) |  | 3295 (52) | 133 (47.9) |  | 3037 (50.5) | 391 (59.5) |  |
| Ecological area |  |  |  |  |  |  |  |  |  |  |  |  |  |
| Northern mountains | 1207 (15.5) | 807 (13.9) | 296 (33.8) | * | 960 (15.3) | 155 (26.4) | * | 1067 (16.3) | 38 (17.4) |  | 1053 (17.9) | 52 (7.1) | ** |
| Red River Delta | 1395 (26.6) | 1187 (29.1) | 111 (19.5) |  | 1199 (28.7) | 103 (21.7) |  | 1244 (28.1) | 60 (27.2) |  | 1177 (28.0) | 127 (28.7) |  |
| North Central and Central Coastal | 1466 (22.4) | 1147 (22.4) | 159 (24.8) |  | 1202 (22.8) | 106 (21.7) |  | 1261 (22.7) | 46 (21.9) |  | 1179 (23.4) | 128 (18.6) |  |
| Central Highlands | 1157 (6.1) | 713 (4.8) | 156 (7.9) |  | 732 (4.7) | 137 (8.9) |  | 812 (5.0) | 55 (7.7) |  | 813 (5.4) | 54 (3.4) |  |
| Southeast | 1218 (15.2) | 941 (14.6) | 55 (4.8) |  | 955 (14.1) | 36 (5.1) |  | 961 (13.4) | 24 (9.4) |  | 833 (11.7) | 152 (22.1) |  |
| Mekong River Delta | 1386 (14.3) | 1122 (15.3) | 102 (9.2) |  | 1123 (14.3) | 105 (16.2) |  | 1170 (14.4) | 52 (16.3) |  | 1046 (13.5) | 176 (20) |  |
| Area of residence |  |  |  |  |  |  |  |  |  |  |  |  |  |
| Urban | 2540 (30.1) | 2005 (31.2) | 190 (17.4) | * | 2020 (30.4) | 175 (20.7) | * | 2103 (29.8) | 86 (22.3) |  | 1928 (27.8) | 261 (39.2) | * |
| Rural | 5289 (69.9) | 3912 (68.8) | 689 (82.6) |  | 4151 (69.6) | 467 (79.3) |  | 4412 (70.2) | 189 (77.7) |  | 4173 (72.2) | 428 (60.8) |  |
| Ethnicity |  |  |  |  |  |  |  |  |  |  |  |  |  |
| Kinh | 6419 (85.0) | 5160 (89.3) | 494 (58.5) | *** | 5260 (87.7) | 406 (66.0) | *** | 5441 (85.9) | 208 (76.9) |  | 4985 (83.5) | 664 (97.1) | *** |
| Others | 1410 (15.0) | 757 (10.7) | 385 (41.5) |  | 911 (12.3) | 236 (34.0) |  | 1074 (14.1) | 67 (23.1) |  | 1116 (16.5) | 25 (2.9) |  |
| Wealth index |  |  |  |  |  |  |  |  |  |  |  |  |  |
| Poorest (Q1) | 1074 (13.6) | 632 (10.6) | 294 (34.5) | *** | 738 (11.7) | 196 (32.2) | *** | 878 (13.5) | 53 (20.8) |  | 895 (15.2) | 36 (6.0) | * |
| Poorer (Q2) | 1735 (16.3) | 1258 (15.2) | 228 (19.5) |  | 1327 (15.3) | 161 (19.1) |  | 1407 (15.4) | 73 (21.3) |  | 1320 (15.3) | 160 (17.8) |  |
| Middle (Q3) | 2029 (22.6) | 1536 (22.9) | 150 (17.1) |  | 1571 (22.7) | 118 (17.1) |  | 1634 (22.3) | 54 (18.4) |  | 1507 (22.0) | 181 (23.0) |  |
| Richer (Q4) | 1981 (30.2) | 1599 (31.3) | 151 (20.4) |  | 1630 (30.7) | 122 (22.3) |  | 1680 (29.9) | 63 (26.8) |  | 1551 (29.4) | 192 (31.7) |  |
| Richest (Q5) | 1010 (17.3) | 892 (20.0) | 56 (8.6) |  | 905 (19.6) | 45 (9.3) |  | 916 (18.8) | 32 (12.8) |  | 828 (18.1) | 120 (21.6) |  |
| **Anthropometric parameters** | | | | | | | | | | | | | |
| Weight (kg) | 19 ± 12.5 | 19 ± 8.9 | 14.5 ± 6 | *** | 18.9 ± 8.4 | 14.3 ± 3.7 | *** | 18.6 ± 8.5 | 15.5 ± 4.4 | *** | 16.3 ± 5.8 | 31 ± 8.7 | *** |
| Height (cm) | 105.5 ± 20.1 | 106.2 ± 19.2 | 96.1 ± 15.3 | *** | 105.4 ± 19.9 | 100.9 ± 15.1 | *** | 104.9 ± 19.6 | 109.5 ± 16.6 | * | 102.2 ± 18.8 | 121.9 ± 14.5 | *** |
| HAZ | -0.65 ± 1.28 | -0.35 ± 1.05 | -2.72 ± 0.70 | *** | -0.45 ± 1.16 | -2.39 ± 0.96 | *** | -0.64 ± 1.28 | -1.07 ± 1.13 | *** | -0.81 ± 1.24 | 0.30 ± 1.05 | *** |
| WAZ | -0.40 ± 1.42 | -0.18 ± 1.28 | -2.03 ± 0.89 | *** | -0.14 ± 1.25 | -2.65 ± 0.59 | *** | -0.32 ± 1.37 | -2.25 ± 0.86 | *** | -0.77 ± 1.11 | 1.75 ± 0.95 | *** |
| WHZ | -0.08 ± 1.14 | -0.04 ± 1.15 | -0.4 ± 0.99 | *** | 0.03 ± 1.08 | -1.41 ± 0.86 | *** | -0.01 ± 1.07 | -2.55 ± 0.52 | *** | -0.23 ± 0.95 | 2.68 ± 0.78 | *** |
| BAZ | 0.01 ± 1.34 | 0.03 ± 1.33 | -0.33 ± 1.07 | *** | 0.15 ± 1.25 | -1.42 ± 0.98 | *** | 0.11 ± 1.23 | -2.53 ± 0.54 | *** | -0.39 ± 1.01 | 2.21 ± 0.69 | *** |
| BMI | 16.3 ± 4.0 | 16.3 ± 3.9 | 15.3 ± 4.2 | ** | 16.3 ± 2.8 | 13.3 ± 1.0 | *** | 16.3 ± 2.7 | 12.6 ± 0.7 | *** | 14.9 ± 1.4 | 20.3 ± 2.3 | *** |
| **Blood biomarkers** | |  |  |  |  |  |  |  |  |  |  |  |  |
| Haemoglobin (g/dL) | 12.3 ± 1.2 | 12.3 ± 1.2 | 12.1 ± 1.3 |  | 12.3 ± 1.2 | 12.3 ± 1.1 |  | 12.3 ± 1.2 | 12.4 ± 1 |  | 12.2 ± 1.2 | 12.6 ± 1 | *** |
| Serum ferritin (µg/L) | 39.7 ± 1.3 | 40.5 ± 1.4 | 38.4 ± 2.8 |  | 39.5 ± 1.4 | 45.5 ± 2.7 | * | 39.5 ± 1.4 | 52.9 ± 4.7 | ** | 38.2 ± 1.4 | 54.2 ± 2.0 | *** |
| sTfR (mg/L) | 6.2 ± 2.6 | 6.1 ± 2.4 | 6.5 ± 3.3 | * | 6.2 ± 2.6 | 6 ± 2.5 |  | 6.2 ± 2.6 | 5.7 ± 1.8 | ** | 6.2 ± 2.7 | 5.9 ± 1.7 | ** |
| Body iron store | 5.5 ± 3.9 | 5.6 ± 3.8 | 5.3 ± 4.1 |  | 5.5 ± 3.9 | 6.1 ± 3.5 | * | 5.5 ± 3.9 | 6.7 ± 2.8 | ** | 5.3 ± 4.0 | 6.8 ± 2.9 | *** |
| Serum zinc (µmol/L) | 9.9 ± 6.3 | 10 ± 6.3 | 8.9 ± 5.2 | ** | 9.9 ± 6.2 | 9.1 ± 5.6 | * | 9.9 ± 6.2 | 8.9 ± 4.4 | * | 9.8 ± 6.1 | 10.8 ± 6.7 |  |
| Serum retinol (µmol/L) | 1.2 ± 0.4 | 1.2 ± 0.4 | 1.1 ± 0.4 |  | 1.2 ± 0.4 | 1.1 ± 0.4 |  | 1.2 ± 0.4 | 1.1 ± 0.4 |  | 1.1 ± 0.4 | 1.3 ± 0.3 | *** |
| RBP (µmol/L) | 1.1 ± 0.4 | 1.1 ± 0.4 | 1.1 ± 0.4 |  | 1.1 ± 0.4 | 1.1 ± 0.4 |  | 1.1 ± 0.4 | 1.1 ± 0.5 |  | 1.1 ± 0.4 | 1.3 ± 0.3 | *** |
| CRP (mg/L) | 1.6 ± 4.5 | 1.6 ± 4.3 | 1.9 ± 5.0 |  | 1.6 ± 4.3 | 1.7 ± 4.4 |  | 1.6 ± 4.3 | 1.5 ± 4.0 |  | 1.1 ± 0.4 | 1.2 ± 0.4 | *** |
| AGP (mg/L) | 0.8 ± 0.3 | 0.8 ± 0.3 | 0.8 ± 0.4 |  | 0.8 ± 0.3 | 0.8 ± 0.3 |  | 0.8 ± 0.3 | 0.8 ± 0.3 |  | 1.5 ± 4.3 | 2.2 ± 4.2 | ** |

AGP, α-1-acid glycoprotein; CRP, C-reactive protein; RBP, Retinol-binding protein; sTfR, Serum transferrin receptor.

Continuous and categorical variables were reported as mean ± SD, and number (percentage), respectively.

P-value was based on t-test and Chi Square test for association for continuous and categorical variables, respectively. * p<0.05; ** p<0.01; *** p<0.001.

Concentration of serum ferritin (µg/L) was presented as geometric mean and standard error (SE).
